# Transgender identity and mental health in adolescence: a scoping review

**DOI:** 10.1101/2020.08.20.20178897

**Authors:** Kirsten L. Patrick

## Abstract

**Background:** Global health guidance has identified gender minorities and adolescents, respectively, as being at elevated risk of mental disorders. The aim of this systematic scoping review was to examine the association between mental distress and transgender status in adolescents, to reflect on how global policy might specifically address the mental health of transgender adolescents.

**Methods:** A systematic search was conducted in six databases – Medline, Embase, CINAHL Plus, ADOLEC, PsychINFO and PsychEXTRA – for published quantitative and qualitative studies examining a range of mental disorders, suicidality and non-suicidal self-injury (NSSI) among adolescents with gender dysphoria or who identify as transgender. The search was limited to original research studies published in Afrikaans, Dutch, English, French and Spanish, but not limited by date. Studies’ prevalence estimates of mental disorders, suicidality and NSSI were abstracted. Meyer’s minority stress model was used as a framework to map risk and protective factors associated with mental distress, grouping by distal stressors, proximal stressors and resilience factors.

**Results:** 49 studies met inclusion criteria. 33 observational studies reported prevalence of depression, anxiety, suicide attempt, suicidal ideation, self-harm, eating disorder and/or disorders of neurodiversity among transgender adolescents. Rates of mental distress were higher among transgender than among both cisgender heterosexual and sexual minority adolescents. Identified risk factors for mental distress were bullying; physical and sexual violence; poor relationships with family and peers; stigmatization by health care providers; internalized transphobia; negative self-concept; and substance use. Factors that appeared to protect against distress included parent-connectedness; peer- and teacher-support; safe school environment; consistent use of chosen names and pronouns; hobbies; and gender-affirming medical treatment.

**Conclusions:** Evidence-informed global governance on adolescent health should adopt a comprehensive, integrated approach to promoting mental health among gender minority adolescents.

**Methods:** A systematic search was conducted in six databases – Medline, Embase, CINAHL Plus, ADOLEC, PsychINFO and PsychEXTRA – for published quantitative and qualitative studies examining a range of mental disorders, suicidality and non-suicidal self-injury (NSSI) among adolescents with gender dysphoria or who identify as transgender. The search was limited to original research studies published in Afrikaans, Dutch, English, French and Spanish, but not limited by date. Studies’ prevalence estimates of mental disorders, suicidality and NSSI were abstracted. Meyer’s minority stress model was used as a framework to map risk and protective factors associated with mental distress, grouping by distal and proximal stressors, and resilience factors.

**Conclusions:** Evidence-informed global governance on adolescent health could encourage countries both to strengthen information systems to support research on transgender adolescent health and to adopt a comprehensive, integrated approach to promoting mental health among gender minority adolescents.

## 1. INTRODUCTION

The 2018 report of the Lancet Commission on global mental health and sustainable development underscored the importance of supporting mental health across the life course, particularly by addressing “social and environmental determinants that…influence mental health at developmentally sensitive periods” such as adolescence and childhood. (1) The Commission noted that specific marginalized population groups are at increased risk of experiencing mental health problems, including transgender people and “young people in vulnerable circumstances”. (1) However, the report did not consider intersectionality of risk and acknowledge the potential overlap of these two groups or the possibility that youth who identify as transgender (that is, as the gender opposite to the gender they were assigned at birth) may be at particularly increased risk of poor mental health.

Adolescents are defined by the World Health Organization (WHO) as individuals aged between 10 and 19 years (early adolescence 10–14 years; late adolescence 15–19 years). (2) Prevalence of self-reported transgender identity among North American adolescent populations ranged from 0.7% to 3.3% in recent studies conducted in different jurisdictions. (3-8) 1.2% of high school students in New Zealand identified as transgender in a 2014 population-based study. (9) 1.6% of German 10- to 16-year-olds were coded as being incongruent to their gender assigned at birth in a recent nationally representative study. (10) Demand for assessment for gender reassignment services in youth appears to be increasing in several countries. (11-14)

While it is well known that sexual minority (gay, lesbian and bisexual – LGB) status is a risk factor for both poor mental health and its determinants, (15,16) less is known about the relationship between mental health and gender minority (transgender, gender diverse and nonbinary – TGDNB) status. A 2011 report from the Institute of Medicine (IOM) in the United States (U.S.) acknowledged that most health research that involved gender minority participants included them with sexual minorities under the umbrella term lesbian, gay, bisexual and transgender (LGBT), and that very little research focused on transgender people exclusively. (17) The IOM report called particularly for research into the health of adolescent and older transgender people. A 2016 review of mental health in transgender youth found only 13 studies that reported on the burden of mental health conditions among children and young people who questioned their gender identity, and limited evidence regarding interventions to address mental distress for this group. (18) Its authors called for more research aimed at understanding the determinants of mental distress among transgender youth and evaluation of interventions to enhance their mental wellbeing.

Current global guidance on adolescent health does not appear to advise countries on the potential for poor mental health among adolescents who identify as a gender other than their assigned gender at birth. The multi-agency Global Accelerated Action for the Health of Adolescents (AA-HA!) Guidance intended to help countries to implement adolescent health strategies (19), the United Nations global strategy for women’s, children’s and adolescents’ health (2016–2030), (20) and WHO recommendations on adolescent sexual and reproductive health and rights, (21) scarcely address the issue of mental health challenges faced by transgender adolescents. Recently updated United Nations technical guidance on sexuality education, however, does discuss the potential health impacts of adolescent gender diversity, acknowledging that “students who are perceived not to conform to prevailing sexual and gender norms, including those who are…transgender”, are vulnerable to violence, and are, if not adequately supported in embracing their preferred gender identity, at increased risk of developing mental distress. (22)

Shifts occurring at the level of global health governance regarding transgender health include adoption of a person-centred, “equity-focused and rights-based” approach to health for transgender people. (23) A 2015 WHO report on sexual health human rights and the law acknowledged the importance to health of being able to express one’s gender identity without stigma, discrimination, exclusion and violence. (24) However, its authors considered adolescents as a separate vulnerable population facing discrimination due to their age; they did not specifically consider the rights of gender minority adolescents.

This review was undertaken to map and narratively synthesize evidence of associations between transgender status and mental distress in adolescents, as well as factors that may protect against mental distress and interventions aimed at enhancing mental wellbeing for transgender adolescents. The purpose was to reflect on how global guidance on adolescent health might support countries to protect and bolster the mental health of transgender adolescents.

## 2. BACKGROUND

A study of the relationship between gender incongruence and mental health among adolescents should consider the context of recent global health policy advances in the areas of adolescent health, mental health and transgender health (see figure 1).

**Figure 1:**
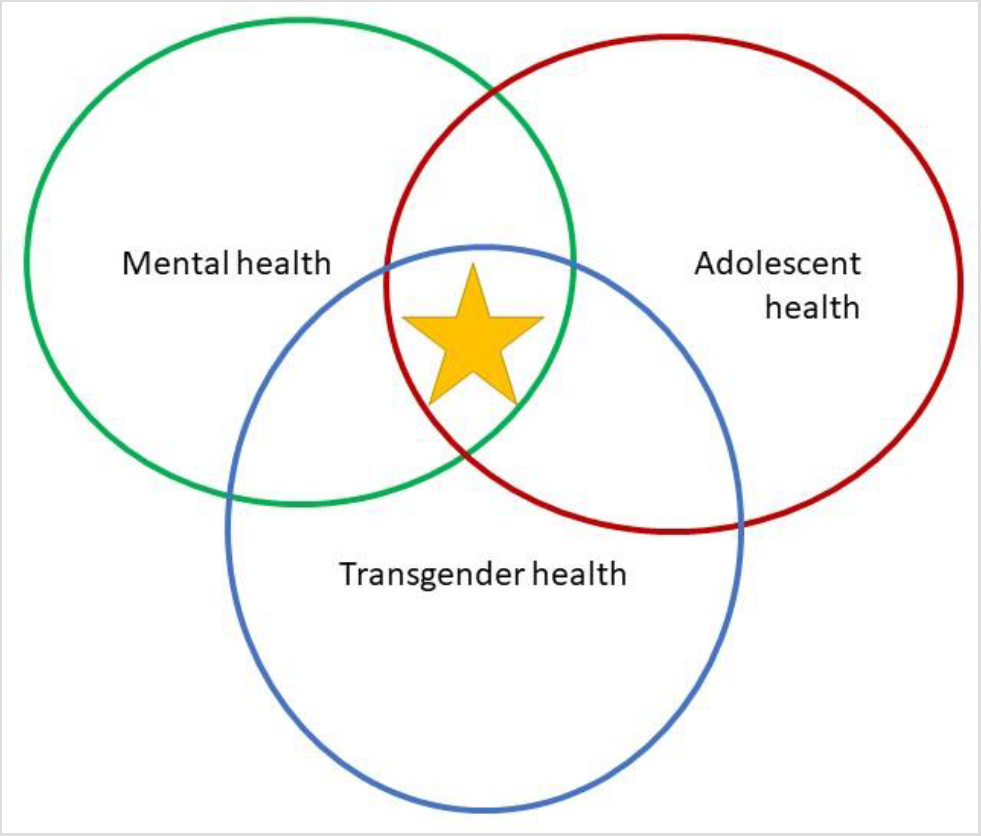
Focus of the project – the intersection of adolescent health, mental health and transgender health

### 2.1 Advances in understanding of adolescent health

The 2016 Lancet commission on adolescent health and wellbeing drew attention to adolescence as a “critical phase in life for achieving human potential…characterized by dynamic brain development” during which individuals develop cognitive, emotional and physical resources necessary for future health, a process greatly influenced by their social environment. (2) “Social adversity” during this critical developmental stage is likely to affect neurodevelopment and reduce adolescents’ current and future capacity to be healthy. (2)

Mental health conditions such as anxiety disorders, depressive disorders and self-harm feature among the top five preventable causes of death for adolescents worldwide, particularly among older adolescents (19) and rank among the top five contributors to loss of disability adjusted life years, both in high income countries (HICs) and in low-and-middle-income countries (LMICs) in 6 of 7 WHO regions. (2)

While the 2016 Lancet commission noted many unique challenges to adolescents’ health globally, it highlighted particularly that socially marginalized groups of adolescents are more likely to experience risks to health and, moreover, that “inequalities established by young adulthood persist and account for many of the disparities in health…and wellbeing in later life.” (2) Among the vulnerable adolescents the AA-HA! guidance identifies are those who are “stigmatized and marginalized because of sexual orientation, gender identity or ethnicity,” those who experience violence within their family or society, and those who are removed from their families or homeless (and therefore have reduced access to health and social services). (19) For some adolescents, however, these vulnerabilities intersect. Such intersectional stigma is likely to exacerbate their risk of poor health, although its effect is difficult to quantify as research in this area is scarce. (25) Adolescent health remains poorly monitored globally due to patchy and inadequate information-gathering and poor disaggregation of data, (19) and monitoring of intersecting vulnerabilities is rare (25).

Adolescents, particularly marginalized ones, also face barriers to accessing health care, not least due to judgement and misunderstanding on the part of health care personnel and regional customs or religious laws that undermine the rights of adolescents to access the health care they need. (2)

### 2.2 Social determinants of mental health and the role of stigma

The Global Burden of Disease Studies of 1990 and 2010 spotlighted a previously under-recognized contribution of mental health disorders to the overall burden of disease, and commentators have emphasized the importance of acting to close the gap between the need for mental health services and their provision. (26,27) However, accumulating evidence has shown that mental disorders are largely socially determined by a range of socio-cultural, political, economic, demographic and environmental factors that interact in a complex fashion from pre-conception to old age. (28) To the extent that people are inequitably affected by these determinants, they are at inequitable risk for mental distress over their lifetimes. (29)

Petersen et al. note that positive mental health encompasses “a subjective sense of one’s own self-worth and the worth of others,… [as] a resource for living”. (30) By extension, a lack of a sense of self-worth or worthiness would impair an individual’s ability to experience mental health. One force that undermines the perceived worth of individuals and groups is stigma, and some evidence links structural forms of stigma to poorer mental health outcomes. (31) Link and Phelan conceptualized stigma as persistent social processes of labelling, stereotyping and othering, which can frequently lead to discrimination of stigmatized people mediated by use of social, economic and political power. (32) Discrimination due to stigma has been termed ‘enacted’ stigma, while impaired self-worth or anticipated rejection resulting from experienced stigma may be considered internalized or ‘felt’ stigma. (33) Even when there is inherent resilience, the effort required to cope with stigma over time may diminish an individual’s psychological resources, which may have negative consequences for both their mental and their physical health, according to Hatzenbuehler et al. (31)

In response to overwhelming evidence of higher observed rates of mental disorders among gay, lesbian and bisexual people compared to heterosexual people, Meyer developed the minority stress theory in 2003 to account for the contributions of stigma –both enacted and internalized – to mental health for sexual minorities. (34) Meyer’s theory, which has recently been adapted to the study of health disparities among gender minorities, (35,36) including transgender adolescents, (37) holds that minorities experience unequal health outcomes because of chronic enacted, internalized and structural stigma experienced alongside other social and structural determinants of health. (38) The theory considers that sexual or gender minority status itself does not confer psychological distress that leads to worse health outcomes. Rather, minority stress and inherent resilience to such stress interact to predict mental disorder. (34) Therefore, relevant factors fit into one of three categories: distal, stigma-based stressors (related to the environment in which the person lives); proximal stressors (related to internalized transphobia – that is, persistent negative feelings about one’s own transgender identity); and protective factors that can be either distal or proximal. (34)

### 2.3 Health inequities for transgender people

Research into health and health disparities for gender minorities is in its relative infancy. (17) Prior to the current decade, studies of transgender individuals mainly enrolled transwomen who have sex with men and the focus of research was usually risk factors for HIV acquisition. (39) Transgender people have been studied within broader “LGBTQ” populations but, until recently, seldom as a separate group that makes a clear distinction between sexuality and gender identity. For example, a 2018 meta-analysis that sought to estimate precisely the risk of attempted suicide among LBGT youth, differentiating by minority group, found that only one of 35 studies reported risk for transgender individuals separately. (40)

The act of not conforming to gender norms is often met with societal hostility and rejection, regardless of gender identity. (41) Transgender people may experience persistent rejection, discrimination and active abuse by family, peers and society from an early age. (42) As a consequence, they may experience other social determinants of poor health including homelessness, interpersonal violence, sexual assault, and may exit education early resulting in more limited employment opportunities. (7,43) Accumulating evidence indicates that transgender youth are more likely to use illicit and harmful substances than their cisgender peers, which correlates with, or increases risk for, other factors that threaten mental wellbeing, such as abuse and victimization. (44) Furthermore, transgender people also frequently face discrimination under the law and may experience barriers to accessing health care services at all levels, or they may forego necessary health care due to stigma. (45,46) Regional guidance for the provision of health care to transgender people, noting disparities in access and health outcomes, has called for interventions to address stigma – e.g., through education in schools, the workplace and medical training to change attitudes – as well as interventions to tackle discrimination – e.g., through legal and policy reforms to advance the right to health of transgender people and dismantle discriminatory structures. (45,47) However, even though deciding on an optimal policy approach may require evidence of effect, stigma may historically have impeded both the allocation of resources to study interventions aimed at addressing health disparities for gender minorities and the enrolment of gender minority subjects in such studies.

Until recently, transgender identity was classified by the WHO’s International Statistical Classification of Diseases and Related Health Problems version 10 (ICD-10) as a mental disorder both for children adults. (48) The 2019 update of the classification (ICD-11) sees gender identity-related health now categorized with conditions related to sexual health, in acknowledgement of evidence from several regions indicating that gender diverse identity is not in itself a mental health issue and that, furthermore, classifying it as such leads to barriers to health care access and stigmatization of TGDNB people. (48-50) This research, undertaken among transgender adults to inform policy, underscored that most transgender individuals became aware of their gender identity in childhood or adolescence, that dysphoria related to gender identity was not a universal experience and that experience of stigma had the strongest impact on their mental health and other outcomes. Given the 2018 Lancet Commission’s imperative to adopt a life-course perspective on mental health, (1) it is important to consider the chronic nature of gender minority individuals’ experience of social adversity and stigma. The extent to which long term health outcomes for transgender people can be improved by efforts to influence early environmental and personal factors remains unclear.

## 3. RESEARCH AIM AND OBJECTIVES

### 3.1 Aim

To systematically identify research evidence examining, firstly, the relationship between transgender status and mental health in adolescents, and, secondly, risk or protective factors related to mental distress in this group – to inform discussion on how global guidance might better address mental health of transgender adolescents.

### 3.2 Objectives

1. Systematically map existing quantitative and qualitative research that examines either the burden of mental distress experienced by transgender adolescents, or the relationship between risk and/or protective factors and mental health outcomes among transgender adolescents.
2. Analyse evidence according to the principles of Meyer’s minority stress theory.
3. Critically consider mapped evidence to reflect on how global health guidance could better support countries to optimize the mental health of their adolescent transgender populations.

## 4. METHODS

A systematic scoping review was conducted, informed by best practice guidance on the conduct and reporting of scoping reviews. (51,52) (See Appendix A for a completed Preferred Reporting Items for Systematic Reviews and Meta-Analyses Extension for Scoping Reviews (PRISMA-ScR) checklist.) No protocol was registered or published. The review was designed to identify published research examining the relationship between transgender status and mental distress in adolescents, to answer the following questions:

- What research exists that examines the burden of mental disorders, self-harm and suicidality among adolescents who identify as transgender?
- Which factors have been shown to contribute to or protect against mental distress in this population?
- What specific interventions have been shown to ameliorate mental distress among adolescents who identify as transgender?

Scoping reviews are intended to map the extent of an existing evidence base on a topic, irrespective of its quality, which justifies a wide search. (51)

### 4.1 Search

A single researcher searched 6 databases – Medline, Embase, CINAHL Plus, ADOLEC, PsychINFO and PsychEXTRA – on 8 June 2019 for studies, conference abstracts and theses, published from database inception to search date, that examined one or more of the following among transgender adolescents: 1) association between gender dysphoria or transgender status and mental distress; 2) factors other than transgender status associated with mental distress; and 3) the effects of any intervention on mental health outcomes. Table 1 outlines the keywords related to search concepts.

**Table 1:** Mapping review search concepts and associated synonyms.

| Concept | TRANSGENDER | ADOLESCENT | MENTAL DISORDER |  |
| --- | --- | --- | --- | --- |
| Synonyms/<br>keywords | Transgender | Teen | Mental health | Mood disorder |
|  | Trans | Teenager | Mental wellbeing | Anxiety |
|  | Transsexual | Teenage girl | Mental distress | Depression |
|  | Travesti | Teenage boy | Mental illness | Substance use disorder |
|  | 2-spirit | Youth | Psychopathology | Alcohol use disorder |
|  | Gender dysphoria |  | Suicide | Autism spectrum |
|  | Gender identity disorder |  | Suicidality | disorder |
|  | Gender non-conforming |  | Self-harm | Asperger |
|  | Gender incongruent |  |  | Eating disorder |

Appendix B outlines the search strategy used in one database (Ovid MEDLINE); searches conducted in the other 5 databases were adapted from this search.

Synonyms for ‘transgender’ included the term ‘transsexual’ (in use until recently), ‘travesti’ and ‘2-spirit’ (cultural terms used in the Americas), and the shortened ‘trans’, which is frequently used as an umbrella term for transgender people as well as those who identify as non-binary, gender fluid or questioning their gender identity. The wider group of gender diverse individuals has recently been characterized as ‘TGDNB’ – trans, gender-diverse and non-binary (53); however, this term was not included in the search as it has not yet been widely adopted. The terms ‘gender non-conforming’ and ‘gender incongruent’ were included, after consultation with a librarian, to ensure identification of all studies of adolescents with gender dysphoria seeking to transition. The historical diagnostic term ‘gender identity disorder’ was included to ensure identification of studies from before 2013. The terms ‘gay’, ‘lesbian’, ‘bisexual’, ‘queer’ and ‘LBGTQ’ were not excluded as transgender people were historically researched within wider sexual minority populations.

Synonyms for the term ‘adolescent’ were limited to those that aligned with the WHO definition, therefore, “young people” and “young adult” were not specifically included as key words, as they might lead to too many irrelevant items that included children and adults up to age 25, respectively, being identified.

Searches were limited to articles published in Afrikaans, Dutch, English, French or Spanish, as these are the languages in which the researcher has reading competency, and to ensure identification of studies from parts of the global South.

Retrieved citations were initially screened by title and abstract, by a single researcher, to identify items matching the inclusion and exclusion criteria outlined in Table 2. Notably, the researcher chose to limit the review to studies looking at outcomes that were either accepted diagnoses of mental disorder, or self-reported symptoms consistent with a diagnosis, as well as self-reported suicidality and non-suicidal self-injury (NSSI) in order to narrow the otherwise broad concept of mental distress. Suicidality and NSSI are strongly associated with mental disorders. (54,55)

**Table 2:** Inclusion and exclusion criteria.

|  | INCLUDED | EXCLUDED |
| --- | --- | --- |
| Study Population | Adolescents aged 10-19 years:<br>>50% of study population to be adolescents, OR<br><50% only if results disaggregated by age and adolescents reported separately, OR<br>mean age of study population within adolescent range, | Children <10 or young adults ≥20 years comprising >50% of study population |
|  | Participants with either gender dysphoria or identifying as transgender | Studies in which study population is defined by sexual minority status (i.e. LGBTQ combined) unless data on transgender participants reported separately, that is, disaggregated reporting of results by gender minority status. |
| Study design | Quantitative: Case series, cohort, case-control and cross-sectional observational studies; intervention studies (controlled/ not)<br>Qualitative: interview or focus group studies | Individual case studies |
| Outcomes studied | Mental disorder diagnosis or self-reported symptoms consistent with mental illness<br>Suicidality (encompassing suicidal ideation and previous suicide attempt)<br>Non-suicidal self-injury (NSSI) | Self-reported substance use (not diagnosed as substance use disorder)<br>Physical health condition including sexually transmitted infection and physical outcome of gender reassignment surgery<br>Satisfaction with gender reassignment treatment |

### 4.2 Approach to data analysis

Full texts of the final selection of studies were retrieved and relevant data charted using an Excel data extraction template with 11 fields as outlined in the Joanna Briggs

Institute guidance for conducting systematic scoping reviews (51), which includes information on study design, population, methodology exposure/intervention, outcomes of interest and key findings. Studies were mapped according to mental health outcome/s examined. The quality of studies reporting prevalence of mental disorder, self-harm or suicidality was assessed using a Joanna Briggs Institute tool for assessing within-study risk of bias, developed to enable critical appraisal of prevalence studies for systematic reviews (See Appendix C). (56) Using this tool, studies are rated on 10 criteria and scored 0 to 10. Studies were assessed as low, medium or high quality by scores of 0–3, 4–6 and 7–10, respectively, as in Garrido-Miguel et al’s recent study (57)

The findings of studies that examined risk or protective factors for mental health, as well as those measuring outcomes of interventions, were mapped to the categories of Meyer’s gender minority stress model: “distal stressors”, “proximal stressors” and “resilience factors”. (35,36)

### 4.3 Ethical considerations & funding

An application for ethics approval was submitted through the LSHTM Ethics Online (LEO) system prior to beginning the review (Appendix D). The LSHTM’s Research Governance & Integrity Office determined that the project did not require ethics approval from the MSc Research Ethics Committee (Appendix E). The project was unfunded.

## 5. RESULTS

Figure 2 details the process of study selection. The initial comprehensive search yielded 4432 items. Deduplication removed 1482 items. Screening of title and abstract for the remaining 2950 items led to removal of 2698 items that clearly did not meet inclusion criteria because they: were not original research articles; reported on single cases; were studies unrelated to transgender issues (e.g. the prefix ‘trans’ was applied to a different word root unrelated to gender, e.g., ‘transdiagnostic’); studied outcomes unrelated to mental health (e.g., studies of HIV risk); or clearly studied non-adolescent populations.

**Figure 2:**
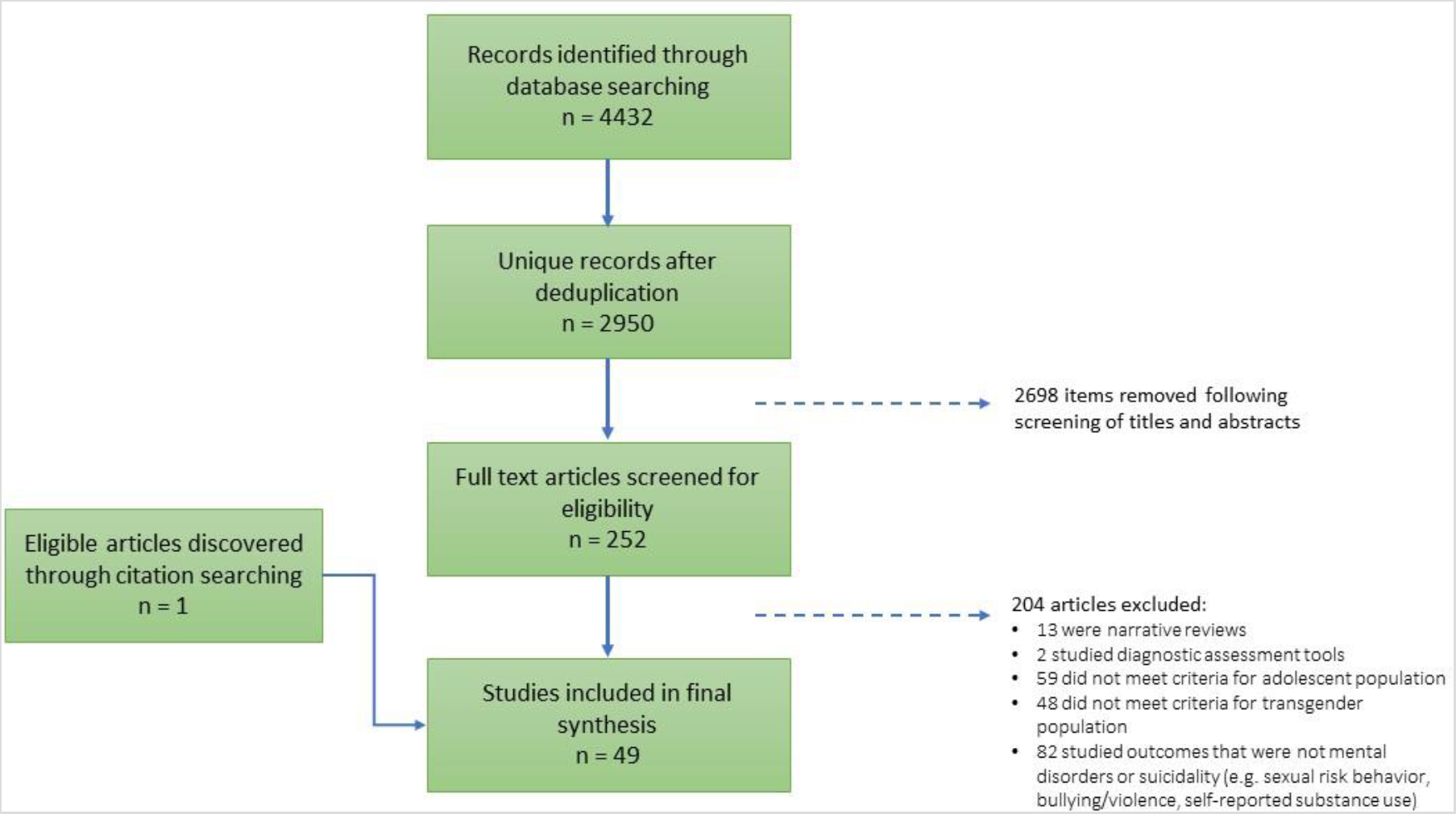
Flow diagram showing study selection

Full texts of 252 articles were retrieved and read by a single researcher. Inclusion and exclusion criteria were carefully applied, resulting in the elimination of a further 204 items (reasons in figure 1). Review of reference lists of full text articles revealed one study not discovered in the initial search. Forty-nine articles (46 original full research studies, 2 published conference abstracts, and 1 national report informed by robust multi-methods research – see Table 3) met criteria for inclusion, 36 of which were published in the last 5 years. Study settings were North America, Europe and Australia.

Sixteen quantitative studies reported on risk factors for mental disorders or mental distress and 3 qualitative studies considered factors that contributed to poor mental health or suicidality. Ten quantitative studies analysed factors that protected against mental distress or promoted mental health, and 2 qualitative studies identified factors that transgender adolescents indicated mental wellbeing. Three uncontrolled studies examined the effect of an intervention on mental health outcomes.

**Table 3.**
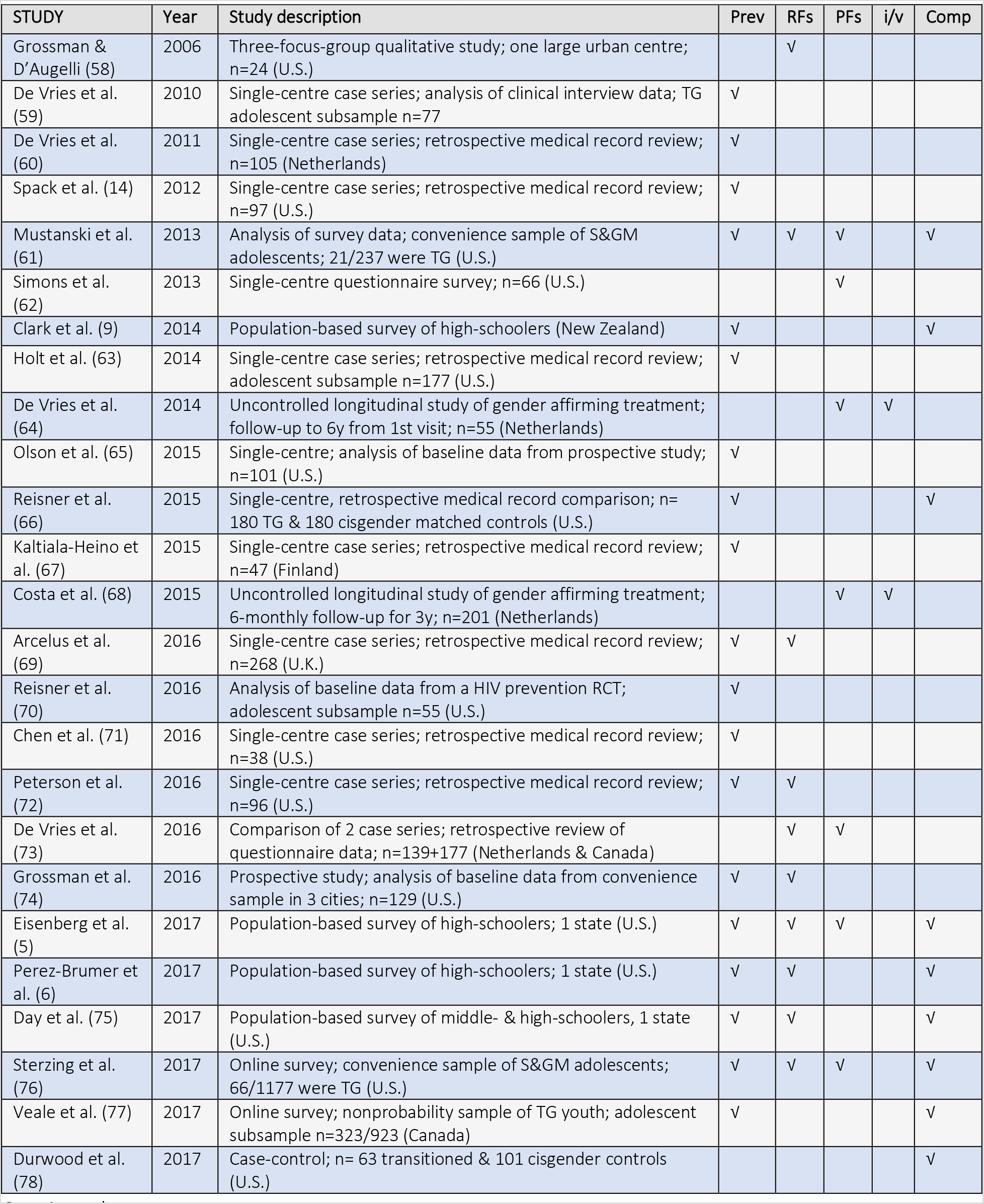
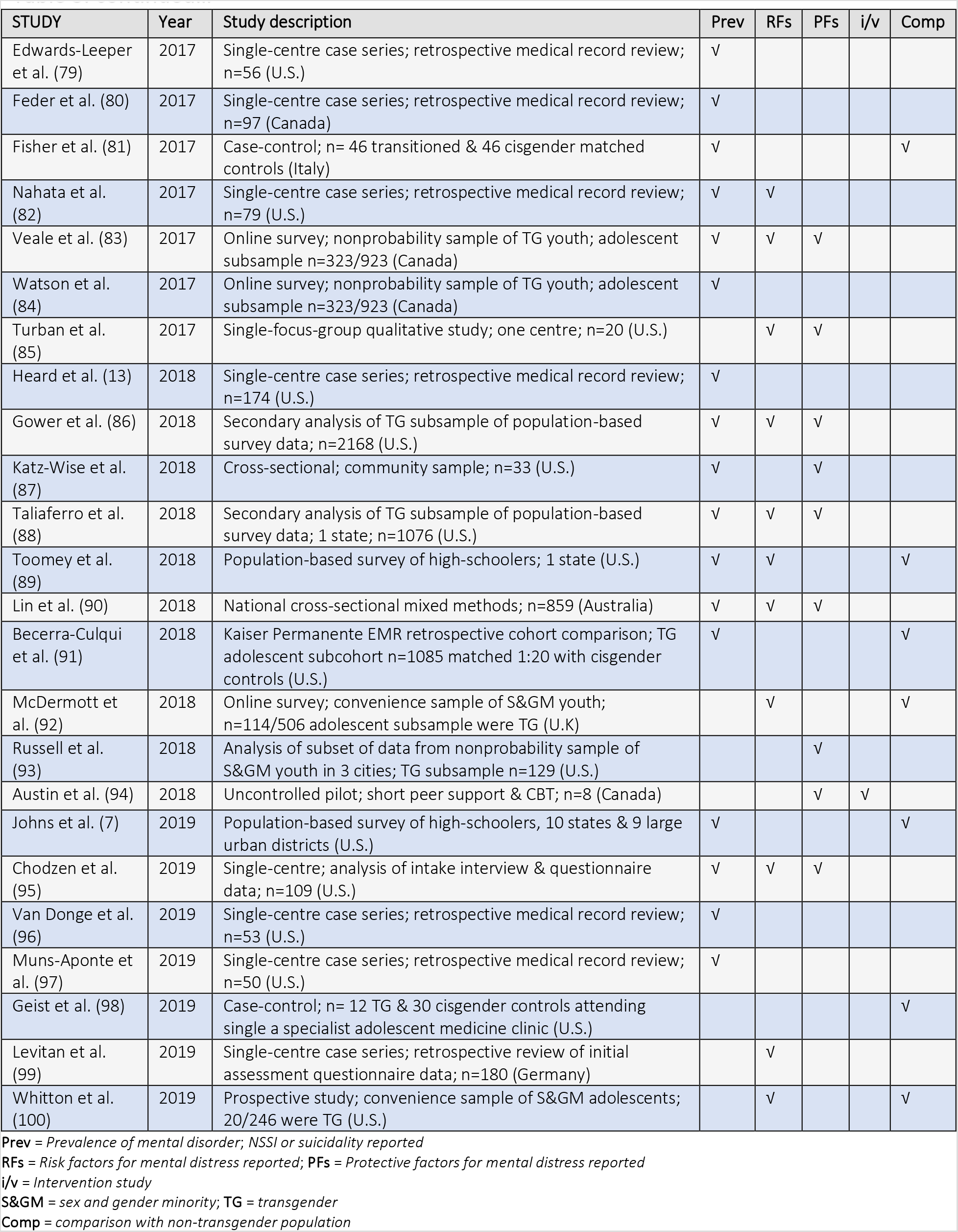
Summary of all included studies (n = 49) of mental distress and associated factors in transgender adolescents, ordered by year of publication.

| STUDY | Year | Study description | Prev | RFs | PFs | i/v | Comp |
| --- | --- | --- | --- | --- | --- | --- | --- |
| Grossman & D'Augelli (58) | 2006 | Three-focus-group qualitative study; one large urban centre; n=24 (U.S.) |  | √ |  |  |  |
| De Vries et al. (59) | 2010 | Single-centre case series; analysis of clinical interview data; TG adolescent subsample n=77 | √ |  |  |  |  |
| De Vries et al. (60) | 2011 | Single-centre case series; retrospective medical record review; n=105 (Netherlands) | √ |  |  |  |  |
| Spack et al. (14) | 2012 | Single-centre case series; retrospective medical record review; n=97 (U.S.) | √ |  |  |  |  |
| Mustanski et al. (61) | 2013 | Analysis of survey data; convenience sample of S&GM adolescents; 21/237 were TG (U.S.) | √ | √ | √ |  | √ |
| Simons et al. (62) | 2013 | Single-centre questionnaire survey; n=66 (U.S.) |  |  | √ |  |  |
| Clark et al. (9) | 2014 | Population-based survey of high-schoolers (New Zealand) | √ |  |  |  | √ |
| Holt et al. (63) | 2014 | Single-centre case series; retrospective medical record review; adolescent subsample n=177 (U.S.) | √ |  |  |  |  |
| De Vries et al. (64) | 2014 | Uncontrolled longitudinal study of gender affirming treatment; follow-up to 6y from 1st visit; n=55 (Netherlands) |  |  | √ | √ |  |
| Olson et al. (65) | 2015 | Single-centre; analysis of baseline data from prospective study; n=101 (U.S.) | √ |  |  |  |  |
| Reisner et al. (66) | 2015 | Single-centre, retrospective medical record comparison; n= 180 TG & 180 cisgender matched controls (U.S.) | √ |  |  |  | √ |
| Kaltiala-Heino et al. (67) | 2015 | Single-centre case series; retrospective medical record review; n=47 (Finland) | √ |  |  |  |  |
| Costa et al. (68) | 2015 | Uncontrolled longitudinal study of gender affirming treatment; 6-monthly follow-up for 3y; n=201 (Netherlands) |  |  | √ | √ |  |
| Arcelus et al. (69) | 2016 | Single-centre case series; retrospective medical record review; n=268 (U.K.) | √ | √ |  |  |  |
| Reisner et al. (70) | 2016 | Analysis of baseline data from a HIV prevention RCT; adolescent subsample n=55 (U.S.) | √ |  |  |  |  |
| Chen et al. (71) | 2016 | Single-centre case series; retrospective medical record review; n=38 (U.S.) | √ |  |  |  |  |
| Peterson et al. (72) | 2016 | Single-centre case series; retrospective medical record review; n=96 (U.S.) | √ | √ |  |  |  |
| De Vries et al. (73) | 2016 | Comparison of 2 case series; retrospective review of questionnaire data; n=139+177 (Netherlands & Canada) |  | √ | √ |  |  |
| Grossman et al. (74) | 2016 | Prospective study; analysis of baseline data from convenience sample in 3 cities; n=129 (U.S.) | √ | √ |  |  |  |
| Eisenberg et al. (5) | 2017 | Population-based survey of high-schoolers; 1 state (U.S.) | √ | √ | √ |  | √ |
| Perez-Brumer et al. (6) | 2017 | Population-based survey of high-schoolers; 1 state (U.S.) | √ | √ |  |  | √ |
| Day et al. (75) | 2017 | Population-based survey of middle- & high-schoolers, 1 state (U.S.) | √ | √ |  |  | √ |
| Sterzing et al. (76) | 2017 | Online survey; convenience sample of S&GM adolescents; 66/1177 were TG (U.S.) | √ | √ | √ |  | √ |
| Veale et al. (77) | 2017 | Online survey; nonprobability sample of TG youth; adolescent subsample n=323/923 (Canada) | √ |  |  |  | √ |
| Durwood et al. (78) | 2017 | Case-control; n= 63 transitioned & 101 cisgender controls (U.S.) |  |  |  |  | √ |
| Edwards-Leeper et al. (79) | 2017 | Single-centre case series; retrospective medical record review; n=56 (U.S.) | ✓ |  |  |  |  |
| Feder et al. (80) | 2017 | Single-centre case series; retrospective medical record review; n=97 (Canada) | ✓ |  |  |  |  |
| Fisher et al. (81) | 2017 | Case-control; n= 46 transitioned & 46 cisgender matched controls (Italy) | ✓ |  |  |  | ✓ |
| Nahata et al. (82) | 2017 | Single-centre case series; retrospective medical record review; n=79 (U.S.) | ✓ | ✓ |  |  |  |
| Veale et al. (83) | 2017 | Online survey; nonprobability sample of TG youth; adolescent subsample n=323/923 (Canada) | ✓ | ✓ | ✓ |  |  |
| Watson et al. (84) | 2017 | Online survey; nonprobability sample of TG youth; adolescent subsample n=323/923 (Canada) | ✓ |  |  |  |  |
| Turban et al. (85) | 2017 | Single-focus-group qualitative study; one centre; n=20 (U.S.) |  | ✓ | ✓ |  |  |
| Heard et al. (13) | 2018 | Single-centre case series; retrospective medical record review; n=174 (U.S.) | ✓ |  |  |  |  |
| Gower et al. (86) | 2018 | Secondary analysis of TG subsample of population-based survey data; n=2168 (U.S.) | ✓ | ✓ | ✓ |  |  |
| Katz-Wise et al. (87) | 2018 | Cross-sectional; community sample; n=33 (U.S.) | ✓ |  | ✓ |  |  |
| Taliaferro et al. (88) | 2018 | Secondary analysis of TG subsample of population-based survey data; 1 state; n=1076 (U.S.) | ✓ | ✓ | ✓ |  |  |
| Toomey et al. (89) | 2018 | Population-based survey of high-schoolers; 1 state (U.S.) | ✓ | ✓ |  |  | ✓ |
| Lin et al. (90) | 2018 | National cross-sectional mixed methods; n=859 (Australia) | ✓ | ✓ | ✓ |  |  |
| Becerra-Culqui et al. (91) | 2018 | Kaiser Permanente EMR retrospective cohort comparison; TG adolescent subcohort n=1085 matched 1:20 with cisgender controls (U.S.) | ✓ |  |  |  | ✓ |
| McDermott et al. (92) | 2018 | Online survey; convenience sample of S&GM youth; n=114/506 adolescent subsample were TG (U.K) |  | ✓ |  |  | ✓ |
| Russell et al. (93) | 2018 | Analysis of subset of data from nonprobability sample of S&GM youth in 3 cities; TG subsample n=129 (U.S.) |  |  | ✓ |  |  |
| Austin et al. (94) | 2018 | Uncontrolled pilot; short peer support & CBT; n=8 (Canada) |  |  | ✓ | ✓ |  |
| Johns et al. (7) | 2019 | Population-based survey of high-schoolers, 10 states & 9 large urban districts (U.S.) | ✓ |  |  |  | ✓ |
| Chodzen et al. (95) | 2019 | Single-centre; analysis of intake interview & questionnaire data; n=109 (U.S.) | ✓ | ✓ | ✓ |  |  |
| Van Donge et al. (96) | 2019 | Single-centre case series; retrospective medical record review; n=53 (U.S.) | ✓ |  |  |  |  |
| Muns-Aponte et al. (97) | 2019 | Single-centre case series; retrospective medical record review; n=50 (U.S.) | ✓ |  |  |  |  |
| Geist et al. (98) | 2019 | Case-control; n= 12 TG & 30 cisgender controls attending single a specialist adolescent medicine clinic (U.S.) |  |  |  |  | ✓ |
| Leviton et al. (99) | 2019 | Single-centre case series; retrospective review of initial assessment questionnaire data; n=180 (Germany) |  | ✓ |  |  |  |
| Whitton et al. (100) | 2019 | Prospective study; convenience sample of S&GM adolescents; 20/246 were TG (U.S.) |  | ✓ |  |  | ✓ |

**Prev** = Prevalence of mental disorder; NSSI or suicidality reported
**RFs** = Risk factors for mental distress reported; **PFs** = Protective factors for mental distress reported
**i/v** = Intervention study
**S&GM** = sex and gender minority; **TG** = transgender
**Comp** = comparison with non-transgender population

### 5.1 Burden of mental distress among transgender adolescents

Thirty-three included studies estimated prevalence of mental disorders, suicidality and non-suicidal self-injury (NSSI) among transgender adolescents. Their findings are summarised in Table 4. All studies but one were judged to be of low or moderate quality according to the instrument used to examine within-study risk of bias. (56)

**Table 4.**
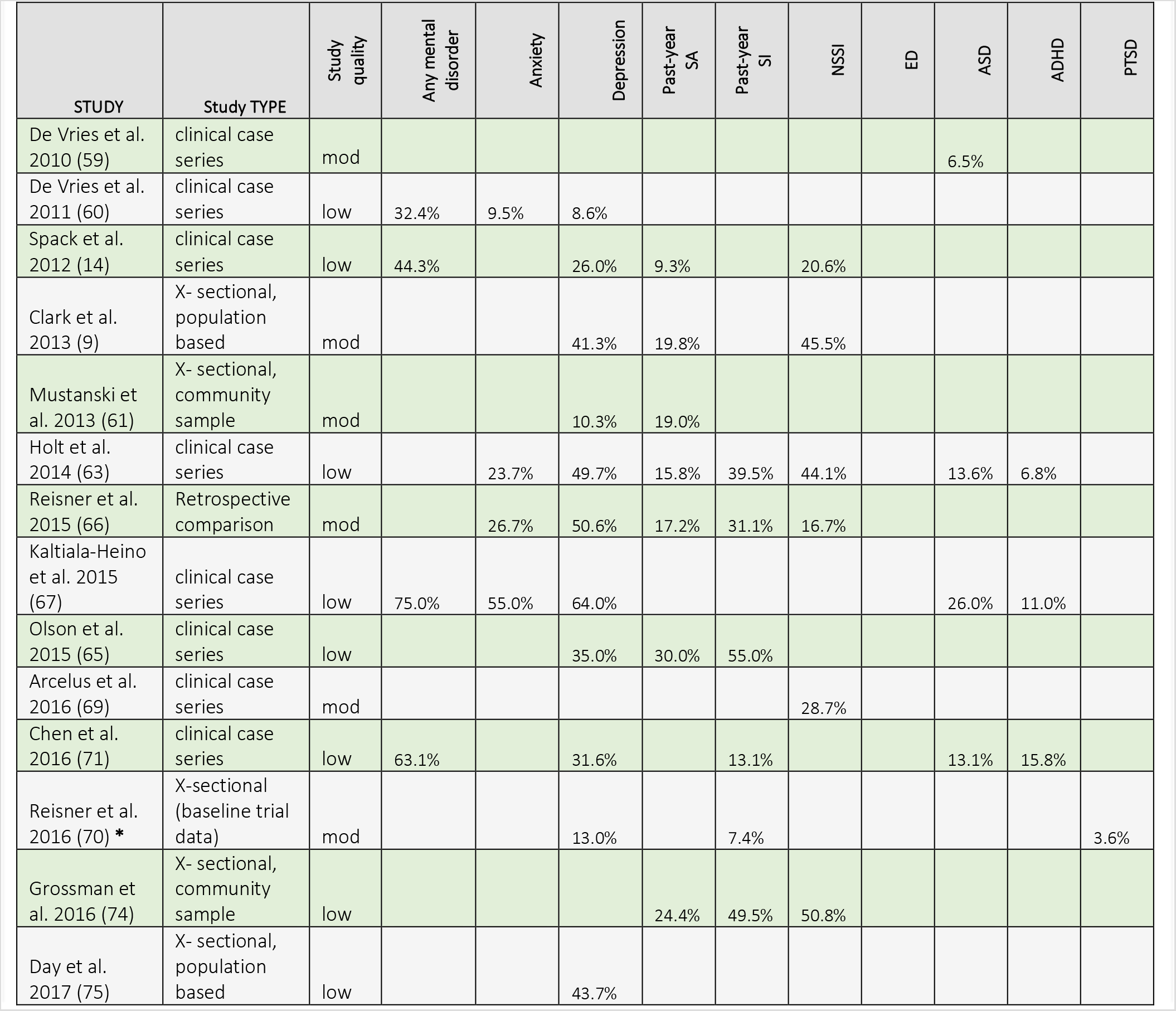
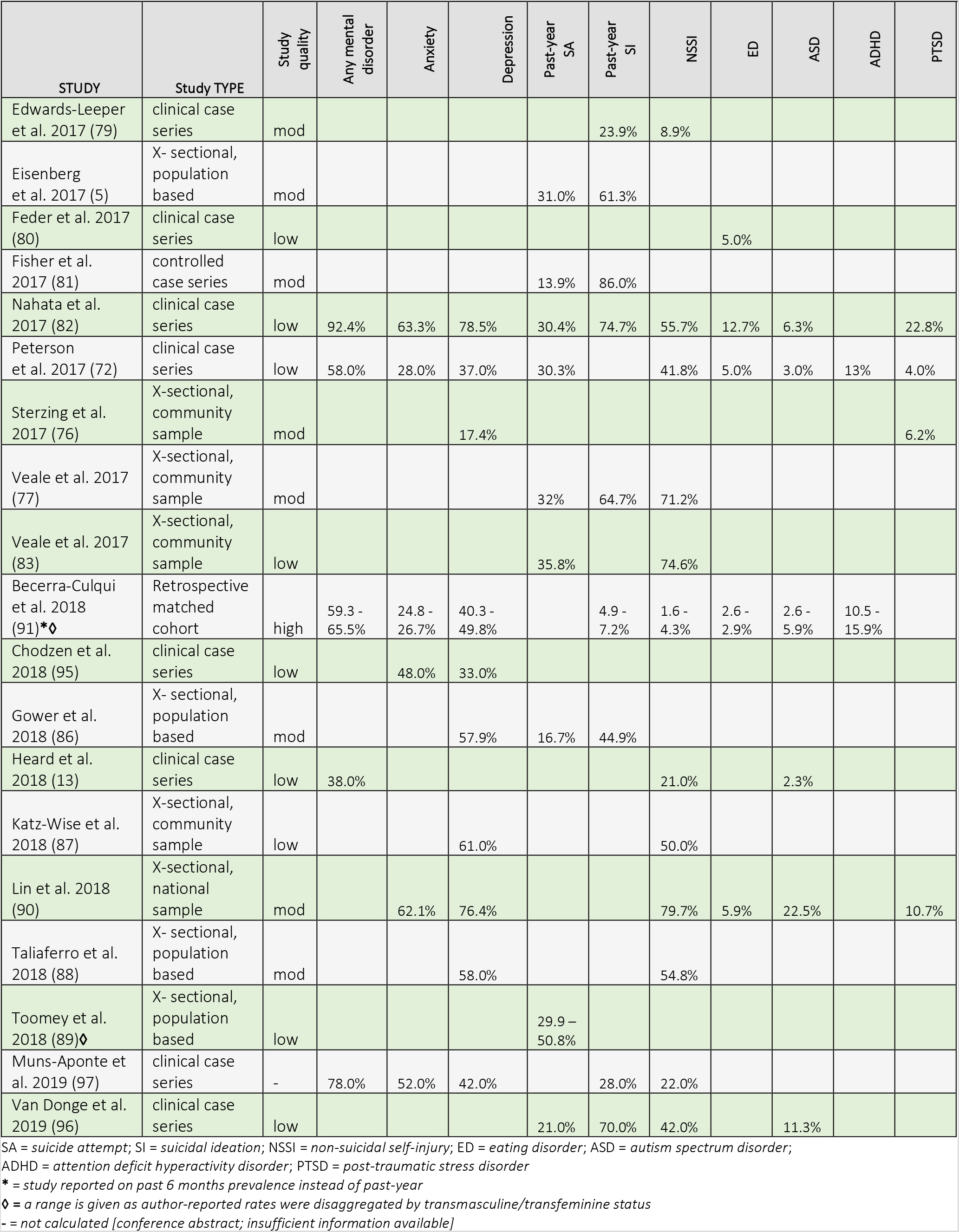
Summary of study estimates of past year prevalence of mental distress outcomes – abstracted from 33 studies.

| STUDY | Study TYPE | Study quality | Any mental disorder | Anxiety | Depression | Past-year SA | Past-year SI | NSSI | ED | ASD | ADHD | PTSD |
| --- | --- | --- | --- | --- | --- | --- | --- | --- | --- | --- | --- | --- |
| De Vries et al. 2010 (59) | clinical case series | mod |  |  |  |  |  |  |  | 6.5% |  |  |
| De Vries et al. 2011 (60) | clinical case series | low | 32.4% | 9.5% | 8.6% |  |  |  |  |  |  |  |
| Spack et al. 2012 (14) | clinical case series | low | 44.3% |  | 26.0% | 9.3% |  | 20.6% |  |  |  |  |
| Clark et al. 2013 (9) | X- sectional, population based | mod |  |  | 41.3% | 19.8% |  | 45.5% |  |  |  |  |
| Mustanski et al. 2013 (61) | X- sectional, community sample | mod |  |  | 10.3% | 19.0% |  |  |  |  |  |  |
| Holt et al. 2014 (63) | clinical case series | low |  | 23.7% | 49.7% | 15.8% | 39.5% | 44.1% |  | 13.6% | 6.8% |  |
| Reisner et al. 2015 (66) | Retrospective comparison | mod |  | 26.7% | 50.6% | 17.2% | 31.1% | 16.7% |  |  |  |  |
| Kaltiala-Heino et al. 2015 (67) | clinical case series | low | 75.0% | 55.0% | 64.0% |  |  |  |  | 26.0% | 11.0% |  |
| Olson et al. 2015 (65) | clinical case series | low |  |  | 35.0% | 30.0% | 55.0% |  |  |  |  |  |
| Arcelus et al. 2016 (69) | clinical case series | mod |  |  |  |  |  | 28.7% |  |  |  |  |
| Chen et al. 2016 (71) | clinical case series | low | 63.1% |  | 31.6% |  | 13.1% |  |  | 13.1% | 15.8% |  |
| Reisner et al. 2016 (70) * | X-sectional (baseline trial data) | mod |  |  | 13.0% |  | 7.4% |  |  |  |  | 3.6% |
| Grossman et al. 2016 (74) | X- sectional, community sample | low |  |  |  | 24.4% | 49.5% | 50.8% |  |  |  |  |
| Day et al. 2017 (75) | X- sectional, population based | low |  |  | 43.7% |  |  |  |  |  |  |  |
| Edwards-Leeper et al. 2017 (79) | clinical case series | mod |  |  |  |  | 23.9% | 8.9% |  |  |  |  |
| Eisenberg et al. 2017 (5) | X- sectional, population based | mod |  |  |  | 31.0% | 61.3% |  |  |  |  |  |
| Feder et al. 2017 (80) | clinical case series | low |  |  |  |  |  |  | 5.0% |  |  |  |
| Fisher et al. 2017 (81) | controlled case series | mod |  |  |  | 13.9% | 86.0% |  |  |  |  |  |
| Nahata et al. 2017 (82) | clinical case series | low | 92.4% | 63.3% | 78.5% | 30.4% | 74.7% | 55.7% | 12.7% | 6.3% |  | 22.8% |
| Peterson et al. 2017 (72) | clinical case series | low | 58.0% | 28.0% | 37.0% | 30.3% |  | 41.8% | 5.0% | 3.0% | 13% | 4.0% |
| Sterzing et al. 2017 (76) | X-sectional, community sample | mod |  |  | 17.4% |  |  |  |  |  |  | 6.2% |
| Veale et al. 2017 (77) | X-sectional, community sample | mod |  |  |  | 32% | 64.7% | 71.2% |  |  |  |  |
| Veale et al. 2017 (83) | X-sectional, community sample | low |  |  |  | 35.8% |  | 74.6% |  |  |  |  |
| Becerra-Culqui et al. 2018 (91)*♦ | Retrospective matched cohort | high | 59.3 - 65.5% | 24.8 - 26.7% | 40.3 - 49.8% |  | 4.9 - 7.2% | 1.6 - 4.3% | 2.6 - 2.9% | 2.6 - 5.9% | 10.5 - 15.9% |  |
| Chodzen et al. 2018 (95) | clinical case series | low |  | 48.0% | 33.0% |  |  |  |  |  |  |  |
| Gower et al. 2018 (86) | X- sectional, population based | mod |  |  | 57.9% | 16.7% | 44.9% |  |  |  |  |  |
| Heard et al. 2018 (13) | clinical case series | low | 38.0% |  |  |  |  | 21.0% |  | 2.3% |  |  |
| Katz-Wise et al. 2018 (87) | X-sectional, community sample | low |  |  | 61.0% |  |  | 50.0% |  |  |  |  |
| Lin et al. 2018 (90) | X-sectional, national sample | mod |  | 62.1% | 76.4% |  |  | 79.7% | 5.9% | 22.5% |  | 10.7% |
| Taliaferro et al. 2018 (88) | X- sectional, population based | mod |  |  | 58.0% |  |  | 54.8% |  |  |  |  |
| Toomey et al. 2018 (89)♦ | X- sectional, population based | low |  |  |  | 29.9 – 50.8% |  |  |  |  |  |  |
| Muns-Aponte et al. 2019 (97) | clinical case series | - | 78.0% | 52.0% | 42.0% |  | 28.0% | 22.0% |  |  |  |  |
| Van Donge et al. 2019 (96) | clinical case series | low |  |  |  | 21.0% | 70.0% | 42.0% |  | 11.3% |  |  |

SA = suicide attempt; SI = suicidal ideation; NSSI = non-suicidal self-injury; ED = eating disorder; ASD = autism spectrum disorder;
ADHD = attention deficit hyperactivity disorder; PTSD = post-traumatic stress disorder
\* = study reported on past 6 months prevalence instead of past-year
♦ = a range is given as author-reported rates were disaggregated by transmasculine/transfeminine status
- = not calculated [conference abstract; insufficient information available]

### 5.2.1 Depression

Twenty-one studies estimated prevalence of depression among transgender adolescents (table 5). Estimates ranged from 8.6% based on clinical diagnosis in small a sample of adolescents receiving treatment to 76.4% based on questionnaire responses in a nationally-representative Australian survey of transgender youth. (60,90) The authors of the Australian study noted that their observed prevalence of moderate to severe depression symptoms among transgender adolescents was almost ten times higher than the 7.7% prevalence observed in a recent national survey of Australian adolescents. (90) A population-based survey of New Zealand high school students found the prevalence of clinically important depressive symptoms among transgender adolescents to be 41.3% compared to 11.8% for non-transgender adolescents. (9) A similar prevalence of depression was observed among transgender adolescents in a population-based survey in California, although the prevalence among non-transgender adolescents was higher at just over 30%. (75)

One study that matched 180 transgender with 180 cisgender older adolescents found a relative risk of diagnosed depression of 3.95 (95% CI 2.60–5.99). (66) Becerra-Culqui et al. reported on a large, retrospective analysis of hospital electronic medical record (EMR) data in 2 states in the U.S., in which 1082 transgender adolescents aged 10–17 years were matched 1:10 with cisgender adolescents. (91) They calculated prevalence ratios (PR) for diagnosed mental disorders and disaggregated their findings by transfeminine vs. transmasculine transgender status. The PR of ever-diagnosis of depression ranged from 4.4 to 7.0, while PR of depression diagnosed in the 6 months before index date ranged from 10.1 to 23.5. That is, these authors found that the odds of a diagnosis of depression in the 6 months before index hospitalization were as much as 23.5 times higher for transgender adolescents than for gender-matched cisgender controls. (91)

A smaller cross-sectional study that compared sexual minority and gender minority adolescents found a higher prevalence of depression among transgender participants than lesbian and gay participants who were cisgender (17.35% vs. 12.8%). (76)

Depression appeared to be more prevalent among older transgender adolescents. Heard et al. found that adolescents who were older at the time of referral for assessment for gender-affirming treatment were more likely to have been diagnosed with anxiety or depression. (13) A similar trend was observed in an analysis of baseline data from a randomised trial of a HIV prevention intervention for adolescent and young adult transwomen, which disaggregated results for adolescents and young adults and found the lowest prevalence of depression for the youngest participants. (70)

### 5.2.2 Anxiety

Ten studies estimated prevalence of anxiety among transgender adolescents (table 5); estimates ranged from 9.5% to 63.3%. Durwood et al. found only slightly elevated rates of anxiety among transgender compared to cisgender young adolescents in their small study comparing 63 socially transitioned younger adolescents with 63 age-matched controls and 38 siblings. (78) However, Reisner et al. observed a relative risk of 3.27 (95% CI 1.80–5.95) for diagnosed anxiety disorder among older transgender compared with cisgender adolescents in a matched analysis of 360 youth. (66) Becerra-Culqui et al. found that for transgender adolescents the odds of ever-diagnosis of anxiety were increased 4- to 5-fold, and the odds of anxiety in the 6 months before consultation for a gender identity issue were increased 8.7- to 18-fold, compared to cisgender controls.(91)

Five cross-sectional studies reported on prevalence of post-traumatic stress disorder (PTSD), which is classified as an anxiety disorder (table 5). One compared transgender and cisgender youth, finding the prevalence of self-reported symptoms suggestive of PTSD to be 6.15% among transgender vs. 4.15% for cisgender adolescents. (76)

### 5.2.3 Self-harm and suicidality

Eighteen studies estimated prevalence of NSSI among transgender adolescents, with estimates ranging widely from 1.6% in a hospital records study to 79.7% in a cross-sectional national survey with convenience sampling (table 5). A Canadian population-based study found a significantly increased risk of past-year NSSI among transgender compared to non-transgender adolescents (prevalence 71.2% vs. 16.5%; relative risk 4.54). (77)

Sixteen studies estimated rates of past-year suicide attempts among transgender adolescents (range 9.3% to 50.8%), and 15 studies estimated rates of past-year suicidal ideation (range 4.9% to 86%) (table 5).

Analysis of data on over 650000 adolescents from two population-based surveys in the U.S. found that suicidal ideation reported by transgender adolescents was twice that of non-transgender peers in one dataset and more than three times higher in the other. (6) Self-reported suicidal ideation among the adolescent subsample within in the recent Canadian Trans Youth Health Survey was found to be 5 times higher than rates reported in comparable population-based surveys of mostly-cisgender adolescents. (77) In a matched cohort comparing transgender and cisgender older adolescents, the relative risk for suicidal ideation was 3.61 (95% CI 2.17–6.03), for a previous suicide attempt 3.2 (95% CI 1.53–6.70), and for past NSSI 4.6 (95% CI 1.95–9.51). (66) However, a small case-control study conducted in a single tertiary care adolescent medicine clinic in the U.S. found no significant difference between transgender and cisgender adolescents with respect to suicidality, self-harm behaviour or depression.(98)

The only study to examine rates of suicidal ideation and NSSI prior to formal gender assessment found that the prevalence ratios for both suicidal ideation and NSSI among transgender adolescents markedly increased in the 6 months prior to gender dysphoria assessment, with up to 54 times the odds for suicidal ideation and up to 143 times the odds for NSSI, compared to cisgender controls. (91)

Comparing sexual with gender minorities, an analysis of data from a cross sectional survey of LGBT adolescents in the UK revealed significantly increased odds of reporting self-harm (OR 1.75; 95% CI 1.09–2.81) and previous suicide attempt or plan (OR 1.63; 95% CI 1.22–2.18) for transgender compared to LGB participants. (92)

### 5.2.3 Eating disorders

Five studies reported on rates of diagnosed eating disorder (ED) among transgender adolescents (table 5), with prevalences ranging from 2.6 to 12.7%. The authors of a large cross sectional study of transgender youth in Canada reported that, among trans adolescents aged 14–18, nearly half reported disordered behaviours related to eating (binge eating or fasting, using pills, laxatives, or vomiting to lose weight) regardless of diagnosed ED, more than twice the prevalence of ED symptoms reported in comparable population-based studies that included mostly cisgender individuals. (84) Eating-disorder-related symptoms are high in cohorts of adolescents with gender dysphoria (72,90) yet rates of confirmed diagnosis are generally much lower (table 5).

### 5.2.3 Disorders of neurodevelopment

Ten studies reported prevalence of autism spectrum disorder (ASD) among transgender adolescents or those seeking to transition (table 5). Rates ranged from 2.3 to 26% across studies. One of the highest rates (22%) of ever-diagnosed ASD was reported in an Australian national cross-sectional study of transgender youth, which also found that 35.2% of participants scored in the range that would warrant further diagnostic tests for ASD on a measure for autism traits, compared to population-based studies that reported such traits in 1.0–2.5% of adolescents. (90) Becerra-Culqui et al. found that prevalence ratios for ASD diagnosis were highest when comparing transfemale adolescents with cisgender females (PR 25.2; 95% CI 12.7 – 52.9). (91)

Five studies estimated prevalence of attention deficit hyperactivity disorder (ADHD) among transgender adolescents (table 5), with range of 6.8 – 15.9% reported. Becerra-Culqui et al estimated moderate prevalence ratios, when comparing transgender with cisgender adolescents (PR 1.3 – 5.3) with respect to ever-diagnosed ADHD. (91)

## 5.3 Analysis of risk & protective factors

Meyer’s minority stress model is used as a framework to present factors found to be associated with mental distress among transgender adolescents. (34) As mentioned previously, the model considers factors in three categories: distal stressors (environmental), proximal stressors (personal) and protective factors (either environmental or personal).

## 5.3.1 Distal stressors

Several studies examined environmental stigma-based stressors experienced by transgender adolescent populations, finding peer victimization, interpersonal violence (IPV) – both physical and sexual – and discriminatory experiences with health care providers to be negatively associated with mental health.

### Bullying victimization and poor peer relationships

A population-based survey of New Zealand high school students showed that 20% of transgender students reported frequent bullying victimization and more than half feared being bullied. (9) Transgender teens were twice as likely to report having been assaulted as their non-transgender peers. Among 47 adolescents who were assessed at one of two adolescent gender identity services in Finland in a 2-year period from 2011 to 2013, 75% of whom were undergoing psychiatric treatment for a non-gender-related psychiatric problem at the time of presentation, 57% reported having been bullied at school and 27% specifically reported gender-identity-related bullying. (67)

A comparison of cross-sectional data on 13- to 18-year-old youth with gender identity disorder seen at gender identity clinics in two countries (Canada – 316 participants – and the Netherlands – 139 participants) found that higher rates of clinically relevant psychological problems in the Canadian sample were associated with poorer peer relationships rather than with demographic factors. (73) Poor peer relationships may indicate victimization.

Among 180 German adolescents with a diagnosis of gender dysphoria, poor peer relations and poor family functioning were found to be significantly associated with clinically relevant behavioural and emotional problems in a cross-sectional study. (99)

Among 2168 transgender adolescents surveyed in a large population-based study of high schoolers in Minnesota, U.S., high rates of NSSI were associated with experiences of victimization. (88) Just over half of transgender adolescents in this subsample had experienced gender-based bullying victimization, and 10% had been physically bullied. Logistic regression analyses revealed that NSSI was significantly associated with depression and any bullying victimization. A U.K. survey of LGBT adolescents found that participants who had experienced gender-based victimization had greater odds of previously planning or attempting suicide than those who were not victimized on the basis of gender expression. (92)

Analysis of data on over 650000 adolescents from two population-based surveys in the U.S. found that peer victimization accounted for 14.7% of the association between transgender status and suicidal ideation, and self-reported depression for almost 18%, suggesting that other risk factors also contribute. (6)

Exploring elements of minority stress as predictors of or protectors against past-year suicidality and NSSI in a large cross-sectional study of transgender youth in Canada, Veale et al. showed that, among 323 trans adolescents aged 14–18, experiences of discrimination were common, with participants experiencing on average 3 kinds of discrimination in the past year. (83) Forms of discrimination included being threatened or physically injured; being sexual harassed, abused or violently assaulted; being verbally victimized for appearance; and being the target of cyberbullying. Scores on an ‘enacted stigma index’ – derived from several measures of discrimination experienced –predicted significantly higher odds of both past-year NSSI and suicide attempt. (83)

Experiencing stigma (as measured using the aforementioned enacted stigma index developed by Veale et al.) was also associated with higher levels of all disordered eating behaviours studied in a large sample of Canadian transgender adolescents. (84)

Qualitative analysis of responses to open-ended survey questions in the Australian national survey of transgender youth revealed that societal experiences of overt discrimination, rejection, deliberate misgendering and other more subtle comments or actions (known as microaggressions) contribute to emotional trauma which drives depression and suicidality. (90) Some adolescents owned that they were stressed by a desire to ‘pass’ as their chosen gender and by fear of being victimized if they failed to pass.

### Intimate partner violence

A prospective study of 248 ethnically diverse sexual and gender minority older adolescents from one urban area in the U.S., who indicated that they had dated, carefully analysed the association between self-reported sexual and/or physical intimate partner violence and a validated global measure of psychological distress. (100) Sexual IPV significantly predicted an increase in psychological distress in within-person repeated measures, suggesting that sexual IPV may contribute to mental illness. In this study transgender adolescents had 2.46 times greater odds of physical partner violence and 3.42 times greater odds of sexual partner violence than cisgender sexual minority youth, suggesting that transgender adolescents have greater exposure to an important risk factor for psychological distress. (100) Substance use, which is also a risk factor for psychological distress, was marginally associated with both forms of partner violence.

### Negative experiences of health care services

A 2006 qualitative, three-focus-group study of transgender adolescents in New York, U.S., explored risk factors for mental distress that participants encountered in the realm of health care. (58) Adolescents reported that unmet mental health service needs, as well as inadequate resources to address their mental health concerns, exacerbated their distress from mental disorders. They also identified as stigmatizing both health care environments that were not affirming and those where they did not feel safe to disclose their gender identity (owing to negative reactions to disclosure in the past). Anticipating such environments sometimes made them avoid seeking health care when they needed it. Negative reactions to disclosure of their gender identity in health care settings could have a severe negative effect on the self-esteem of transgender youth.

A more recent, low quality, single-focus-group study of 20 transgender adolescents sought to understand problems that youth encounter in the health care setting, with the purpose of informing practice improvements. (85) Interviewees highlighted the importance to their mental wellbeing of acceptance of their transgender status by health care providers. They reported that discrimination, judgement and misgendering in the health care environment add to mental distress and lead to lack of engagement with services. “Many of us are anxious and depressed not because we are transgender but because other people have a problem with us being transgender,” noted one participant. (85, page 276) Furthermore, having to shoulder the burden of responsibility for educating providers and ensuring they received an acceptable standard of treatment is stressful and exacerbates mental illness. Participants emphasized that access to gender affirming treatment including medication improves mental wellbeing, reduces body dysmorphia and attenuates gender dysphoria.

In the qualitative analysis of survey responses in the aforementioned Australian study, transgender adolescents with eating disorders reported feeling burdened by having to convince family and health professionals that eating disorders are different from gender dysphoria symptoms (i.e., others assumed that the motivation for undereating or overexercising was merely to control the appearance of secondary sexual characteristics or body shape and not reflective of an eating disorder). (90) Participants said that feeling dismissed by health care providers as just seeking attention added to the mental distress associated with their eating disorder.

## 5.2.2 Proximal stressors

Two included studies sought to understand the relation between high rates of mental illness and person-level factors including internalized transphobia and substance use.

Among 106 transgender and gender nonconforming 12- to 18-year-olds treated at one interdisciplinary gender identity clinic serving an urban region in the U.S., participants who scored high on the ‘internalized transphobia’ domain of the Gender Minority Stress and Resilience Scale were significantly more likely to meet diagnostic criteria for major depressive disorder or generalized anxiety disorder. (95)

An analysis of data on 36070 high school students who participated in a population-based survey in California (1.2% of whom reported transgender identity), depressive symptomatology was shown to be a partial mediator of higher rates of substance use (2.5 to 4 times higher, depending on substance category) among transgender compared to non-transgender adolescents. (75)

## 5.2.3 Protective factors

Several studies identified external and internal factors that appeared to protect against mental disorder or reduced the likelihood of suicidality and self-harm among transgender adolescents, including family- and social-connectedness, safe environments, aspects of affirmative treatment, hobbies and a sense of belonging.

### Personal-level factors

In the cross-sectional survey of Australian transgender adolescents, 83.4% of participants indicated that they used music or art to feel mentally better, and 65.5% said caring for pets improved their mental health. 65% used books/reading, 60% used movies, 44% used group activities and 32.8% used exercise to enhance wellbeing. (90) In responses to open-ended questions, participants reported often using such activities to escape from the realities of their lives. Many used video gaming for the same purpose. One transfemale participant described the effect of video gaming thus: “[It] allow[s] me to enter a space of personal comfort, where I can express myself freely and honestly and for a time ignore the reality of my difficulties. [I can] present as female, playing female characters, for example.” (90, page 72) While substance use was associated in many other studies with poor mental health and risk factors for mental distress, some in the Australian study explained that they used illicit substances as a way to escape from gender dysphoria and mental distress: “Sometimes when I do drugs I forget that I was born female and I think I am male: it’s really nice to have that feeling.” (90, page 74) Some participants remarked that having role models and celebrity representation helped them to be hopeful for their future (a state that may temper suicidality).

### Family and social supports

A secondary analysis of data on 2168 adolescents who identified as transgender in the 2016 Minnesota Student Survey considered the protective potential of 8 factors (connectedness to parents, adult relatives, friends, adults in the community, and teachers; youth development opportunities; and feeling safe in the community and at school) – separately and together – against depression, suicidality and substance use. (86) Each protective factor was significantly associated with lower odds of depression, suicidality and substance use in individual adjusted regression analyses. When all protective factors were considered simultaneously, parent connectedness and feeling safe at school were the significant contributors to reduced depression levels and suicidality. Greater connectedness to parents and non-parental adults was found to significantly attenuate the relationship between bullying experience and any NSSI. Only high parent connectedness and perception of safety in the school environment were significantly associated with reduced repetitive NSSI. High parent-connectedness and high quality of teacher-student relationship, together, were significantly associated with reduced substance use. (86)

Among 66 transgender adolescents assessed at a single gender identity clinic in the U.S., higher perceived parental support (as measured by a validated instrument) was associated with fewer reported symptoms of depression. (62)

Veale et al., in their study of the role of elements of minority stress in predicting NSSI and suicidal ideation, found that higher score on a measure of family connectedness was significantly associated with lower odds of NSSI, while higher score on a tool that measured friends’ caring was associated with lower odds of past-year suicide attempt in multivariate logistic regression models. (83) Furthermore, when protective factors such as family connectedness, school connectedness and friends caring converged, the probability of past-year suicide attempts reduced considerably. Watson et al. found that the social buttresses of strong family support, school connectedness and friends caring attenuated the relationship between stigmatization and disordered eating among Canadian transgender adolescents. (84)

Fifty-four per cent of transgender adolescents surveyed in the Australian national study indicated that they used counselling or mental health services, and 40.2% attended peer-led safe social spaces, to support their mental wellbeing. (90) Qualitative analysis of responses to open-ended questions revealed that youth found social support, from both family and from support groups, to enhance their mental wellbeing owing to its effect on positive self-concept and self-acceptance. However, transgender adolescents often found it difficult to find support *within* the transgender youth community where many people may be experiencing trauma. As one participant noted, “When everyone has varying mental health problems, it makes it more difficult to support one another because in reality we all need support.” (90, page 50)

Austin et al. piloted a group-based intervention (“AFFIRM”) that comprised pyscho-education, interactive learning, group rehearsal and peer support, in an uncontrolled study conducted with 8 transgender adolescents recruited from a Canadian urban centre. (94) The single-weekend intervention reduced participants’ depression scores both immediately and at 3-months follow-up although a meaningful effect on general levels of coping was not seen.

### Use of chosen names and pronouns

Russell et al. studied the effect on depression and suicidality of chosen name use in several contexts among 129 older transgender adolescents who used a name other than their given birth name, selected from population-based samples in three cities in the U.S. (93) Those adolescents who reported that others used their chosen name in more of the studied contexts (home, school, work, with friends) were significantly less likely to report symptoms consistent with a diagnosis of depression, or suicidality, after adjustment for social support. Each additional context in which their chosen name was used predicted a 29% decrease in suicidal ideation and a 56% decrease in overall suicidal behaviour. (93)

*Gender affirming medical treatment*

The Australian national survey of transgender adolescents found that many adolescents indicated that being able to receive puberty-suppressive or gender-affirming hormonal treatment substantially improved their mental wellbeing. (90) According to one transmale participant who responded to open-ended questions, “Being able to have my body slowly changing in a direction that reflects more of how I feel about myself significantly improved my mental health and wellbeing.” (90, page 71)

A prospective study examined psychological outcomes for 201 adolescents with gender dysphoria aged 12 to 17 years, who received guideline-standard psychological support intervention and puberty suppression treatment between 2010 and 2014 at one clinic in the Netherlands. (68) Participants had six-monthly follow up over an average of 17 months, with measurement of psychological and global functioning on validated instruments at 3 time points. Adolescents’ average global functioning improved significantly after 6 months of psychological support, and those who received treatment to suppress puberty at the same time as psychological support had significantly improved psychosocial functioning at 12 months compared with their scores after only 6 months of psychological intervention.

In another prospective study Dutch researchers followed 55 transgender adolescents (mean age 13.6 years at baseline) for 6 years to young adulthood, to assess their global and psychological functioning, depression, anxiety, body image, social and educational functioning and quality of life before and after gender reassignment treatment. (64) All participants reported improved psychological wellbeing after treatment that included puberty suppressant medication and gender-affirming hormone therapy.

## 6. DISCUSSION

### 6.1 Summary of the findings of this review

This scoping review identified 49 studies that either found transgender adolescents to experience a high burden of mental disorders and suicidality or identified factors contributing to mental distress among transgender teens. Rates of mental distress and suicidality were observed to be substantially higher among transgender than both cisgender heterosexual and sexual minority (LGB) adolescents, in some studies.

Furthermore, use of Meyer’s minority stress model (34), adapted for use in gender minorities (35), as a framework to map evidence on risk and protective factors, helped to highlight salient factors affecting mental health among transgender adolescents that may be modifiable by social or health policy interventions. Risk factors for mental disorder and suicidality in this population included enacted non-acceptance (by parents, wider family, peers and health care workers), as well as various forms of victimization (from verbal microaggressions and cyber bullying through to physical and sexual IPV). Factors that appeared to bolster internal resilience, support mental wellbeing and reduce self-harm included social acceptance and affirmation of adolescents’ gender diversity – through strong parental, family and peer support, use of correct names and pronouns supportive school environments and guideline-congruent gender-affirming medical treatment.

### 6.2 What this study adds

A scoping review with a similar question was published in 2016. (18) Connolly et al. had slightly different inclusion criteria; they included studies published in English in 2 databases between January 2011 and March 2016 looking at mental health in individuals with gender dysphoria up to mean age 24. They found only 15 studies, 7 of which did not meet inclusion criteria defined for this review. (18) This review identified 49 studies of transgender adolescents, only 2 of which were published before 2011, and 21 of which were published after the inclusion period of the earlier review. This project, therefore, extends understanding of the burden of mental illness experienced by transgender adolescents and underscores a recent uptick in the volume of research on mental health of transgender adolescents, albeit only in high income countries.

Published evaluations of well-designed interventions to prevent mental illness or promote mental wellness among transgender youth are still relatively few. Only 2 new intervention studies were published since the 2016 review. A large, multi-site prospective, uncontrolled study in the U.S. has completed enrolment and is set to examine, among other outcomes, the long-term mental health outcomes of gender-affirming treatment in 280 pubertal and post-pubertal transgender adolescents. (101)

Mapping of risk and protective factors, using the minority stress model, not only enabled categorization of the main contributing factors, it also revealed that relatively little is known about proximal compared to distal risk and protective factors. Knowing little about the negative thoughts that transgender adolescents direct towards self on account of their transgender identity (internalized transphobia) and the private self-defeating actions they may take as a consequence, could make it difficult to develop interventions targeted towards addressing proximal risk factors. This is important to highlight as it may give direction to future research. The ability of quantitative research designs to identify aspects of internalized transphobia may be limited. Understanding of transgender adolescents’ relevant thoughts and perceptions could be acquired more effectively via well-conducted qualitative studies. The present scoping review found only 3 studies that used qualitative methods, and 2 of these focused mainly on adolescents’ perceptions or experiences of health service access rather than on aspects of internalized transphobia.

### 6.3 Limitations of the review

There are several limitations to this project, which warrant consideration.

Firstly, the review examined gender as a binary construct. The search did not include the terms ‘non-binary’, ‘gender fluid’ ‘questioning’ or ‘TGDNB’ among the synonyms for transgender and gender dysphoria. The conceptualization of gender as binary in this review, although a limitation imposed by the researcher to keep the project within the realm of manageability, may have compromised the richness of the findings. While a few included studies did identify subpopulations of gender minority, the spectrum of gender diversity beyond transgender was not explicitly considered in this project. Recent expert opinion, however, is that researchers and health professionals ought not to conceptualize gender as binary, since such categorization does not match gender minorities’ experience, (102) but rather understand gender as “a spectrum or galaxy” rather than a binary construct where some people transition from male to female or vice versa. (53) Furthermore, it is possible that in some global settings where sex and gender diversity is not culturally or legally accepted, transgender individuals may take a more gender-fluid approach to gender expression, adaptive to the safety of their environment. Therefore, limiting the present review to a more binary concept of gender, may have resulted in studies of gender fluid adolescents being missed, if any exist.

Secondly, some individual participants in studies included in the review were outside the adolescent range as defined by the WHO. This is because not all studies that considered adolescent populations used the WHO definition for adolescents; some included participants up to the age of 24, while others considered both children and adolescents. To avoid excluding studies with valuable information on adolescent participants, a decision was taken to include studies of wider populations **if** the mean age of participants in the study was within the adolescent range. However, rates of symptoms of mental disorders have been shown to differ between adolescents and young adults. For example, two studies included in the review reported findings separately for youth aged 14–18 years old (these data were included in the current review) and 19–25 years old (these data were not included). (83,84) The authors found rates of eating disorders, suicidality and self-harm to be somewhat higher among 14- to 18-year-olds than among 19- to 25-year-olds. Therefore, including studies in which a few participants were outside of the adolescent age range may have influenced estimates of burden of mental distress.

Thirdly, as stated in the methods, a decision was made to limit the review to studies looking at outcomes that were mental disorder diagnoses, suicidality or NSSI in order to clarify more precisely the otherwise broad concept of mental distress. As such, several studies in which the outcome of interest was self-reported substance use were excluded because the adolescent participants were not diagnosed with substance use disorder. However, even moderate substance use may be an indicator of mental distress and substance use has been associated with both mental disorders (103-105) and factors that were discovered in this review to be associated with mental health. (105,106) It is, therefore, possible that this project may have forfeited some depth of understanding related to risk and protective factors associated with mental health among transgender adolescents by excluding studies that looked only at substance use.

Lastly, a single researcher conducted the search, screened identified items, applied inclusion and exclusion criteria and made the final selection of studies, which would have biased the findings of this review. However, the researcher was required to work alone for this project.

### 6.4 Gaps in the evidence base

As all studies included in the review were conducted in HICs, it is difficult to interpret the findings for the global context. While this may be considered a limitation of the search it is more likely a limitation of the evidence base, as six databases were searched for studies in any of five languages. Exclusion of non-English language papers has, furthermore, been shown to have little effect on the findings of systematic reviews of interventions in biomedicine. (107,108) Although a few articles identified in the initial search were in French or Spanish language, none of these met inclusion criteria for the final review. (109,110)

Very few studies included in this review considered the intersection of minority stress and other vulnerabilities such as ethnicity and poverty, which is perhaps unsurprising since the authors of a 2019 methods review described intersectional stigma research as ‘nascent’. (25). Turan et al. defined intersectionality as “a lens through which researchers seek to understand the complex nature of identity, health, social relationships and power that plays out within human interaction and experiences.” Most studies of stigma-related risk factors (including those in the current review) consider one or two factors, yet pathways of stigma and discrimination are complex.

## 7. IMPLICATIONS FOR GLOBAL HEALTH GUIDANCE

A stated objective of this review was to reflect on how global guidance could incorporate the accumulated evidence to support countries in optimizing the mental health of their adolescent transgender populations. As mentioned in the introduction, current global guidance on adolescent health and mental health barely signal awareness of the problem that mental distress among transgender adolescents is remarkably common and that risk factors for mental distress in this population have been identified.

Despite the findings of this review being representative of only a few HICs, studies that have examined mental distress among transgender adults beyond North America, Europe and Australia suggest that gender minorities’ experiences in adolescence may be common across more countries. A recent large cross-sectional study of young transgender adults in 32 Chinese provinces found high rates of lifetime suicidality, self-harm and mental disorders, compared to estimates for the general population, as well as risk factors for mental distress similar to those mapped in this project. (111) These findings chime with those of smaller quantitative and qualitative studies conducted in Turkey and Mexico, and among LGBT refugees to North America from a wide range of countries. (49,112-114)

Yet acknowledging the problem, and allowing that it is likely a global problem, would not be enough. Health policy-makers also need guidance on strategies to address the issue. The findings of this project allow for several policy suggestions, which align well with the main objectives of the WHO’s mental health action plan 2013–2020 (115):

### I. Strengthen effective leadership and governance in the area of transgender adolescent mental health

Deeply held beliefs about gender and sexuality are reinforced globally by education, religion and culture. Moreover, stigma may be enacted through policies supported by law. Many countries still criminalize same-sex relations and some also outlaw forms of gender expression. (116) Adolescent health has quite rightly been prioritized in global health governance. (19) Leveraging the valuable normative role of global guidance on countries’ health policy-making by making global guidance more explicit regarding the specific mental health burdens on transgender adolescents and specific policy actions that may protect their mental health, would ensure that gender minority adolescents are not left behind.

### II. Develop guidance on a comprehensive, integrated approach to promoting mental health and preventing mental distress among transgender adolescents

The 2017 AA-HA! guidance underscored the importance of taking an “Adolescent Health in All Policies (AHiAP)” approach to improving adolescent health at the country level. (19) The AHiAP approach, which considers policies and programs in sectors other than health – such as education, justice and social protection – and their implications for adolescent health and wellbeing, would be a good basis for developing guidance for countries on how to prevent mental distress for gender minority adolescents.

This project mapped many social factors that underpin poor mental health for transgender adolescents. In a review that considered stigma as a fundamental cause of population health inequalities, Hatzenbuehler et al. emphasized that stigma is a powerful and persistent influence on health because its effects are mediated by so many other social determinants. (31) They also noted that acting to reduce health disparities is challenging as stigmatizers retain power differentials with respect to a stigmatized group through several different mechanisms or structures, so that, if the power of one mechanism is reduced, stigma can still be enacted through intensification of other mechanisms. Therefore, taking a multi-level approach to the problem that tackles legal protections, the education system, training of health workers and development of social supports is likely to be the most effective strategy, similar to the approach recommended by authors of a systematic review of interventions to reduce HIV related stigma and discrimination. (117) Complex approaches that may offer good return on investment could include the following elements: online and community interventions that seek to maximize both safety and support for transgender adolescents; school interventions along the lines of Gay-Straight Alliances (106) and training educators as allies (118); interventions that can help parents and families to support transgender adolescents (39); and wider education of health care providers on how to improve access to treatment through gender affirming care (102, 119).

### III. Strengthen information systems and support research on the mental health of gender diverse adolescents

This project found that robust population-based estimates of the prevalence of gender minority status among adolescents are limited to a few HICs. Existing studies that have estimated the burden of mental distress in this population are generally of poor quality and no such studies have been conducted in LMICs. Global guidance could advise countries to track gender minority status in surveys of adolescents or scholars. Linkage with routinely-collected robust measures of mental health and distress, as well as risk factors for mental health, could allow for higher quality research that takes an appropriate intersectional approach to examining the effects of enacted stigma in different jurisdictions. Collection of expanded information on gender identity by health care institutions could also augment research in this area, and support research to move away from a binary concept of gender, as previously suggested. (53,102) As identified above, knowledge about modifiable proximal risk factors for mental distress are lacking and are needed to inform appropriate interventions. Qualitative research to discover modifiable proximal risk factors could be encouraged. Well-conducted evaluations of interventions that seek to address the established drivers of mental distress for transgender adolescents are lacking and could also be encouraged and supported.

## 8. CONCLUSION

Adolescents who identify as transgender are at remarkably high risk of mental disorders, self-harm and suicidality, according accumulating evidence from several jurisdictions internationally. Risk factors that contribute to the high burden of mental distress in this vulnerable population have been identified, but more research is needed to understand the extent to which risk factors may be modifiable. However, the evidence base is compelling enough to warrant stronger guidance from global institutions to encourage member countries both to strengthen information systems to support research on transgender adolescent health and to adopt a comprehensive, integrated approach to promoting mental health among gender minority adolescents. This heretofore overlooked vulnerable subpopulation must not be left behind.

## Data Availability

All data included in this review are published.

## KEY DEFINITIONS AND ABBREVIATIONS USED IN THIS REVIEW

Adolescent: individual aged 10 to 19 years
Transgender: state of identifying as the gender opposite to the gender assigned at birth; sometimes used more expansively to apply to all gender minorities
TGDNC: transgender, gender-diverse and/or non-conforming
Transfemale: female who was assigned male at birth
Transmale: male who was assigned female at birth
Gender minority: person who does not identify with their assigned gender at birth, i.e., person who is trans or TGNDC; person may be same- or opposite-sex attracted
Sexual minority: person who is sexually attracted to a person of the same gender or persons of both genders or not sure, i.e., not heterosexual
LGBTQ: lesbian, gay, bisexual, transgender, questioning individuals (sometimes limited to LGB or LGBT only); umbrella term for sexual and gender minority people
Gender dysphoria: officially a mental disorder defined in the DSM 5; often used more loosely to refer to physical, emotional and social distress experienced by a person who does not identify as their gender assigned at birth and who has not yet transitioned.
Gender nonconformity/ gender incongruence: behaviours and expression of self/person that do not conform with society’s usual expectations for assigned gender; sexual minorities and cisgender people may display gender nonconformity
Suicidality: previous suicide attempt/s and/or suicidal ideation/plans **NSSI** – non-suicidal self-injury (also referred to as self-harm)
Mental distress: for the purposes of this review the term refers to one or more of: a diagnosed mental disorder, self-reported symptoms consistent with a mental disorder, previous suicide attempt, self-reported suicidal ideation or self-reported physical NSSI
ASD: autism spectrum disorder
ADHD: attention deficit hyperactivity disorder
PTSD: post-traumatic stress disorder

## 10. APPENDICES

## APPENDIX A: Copy of the Preferred Reporting Items for Systematic reviews and Meta-Analyses extension for Scoping Reviews (PRISMA-ScR) Checklist

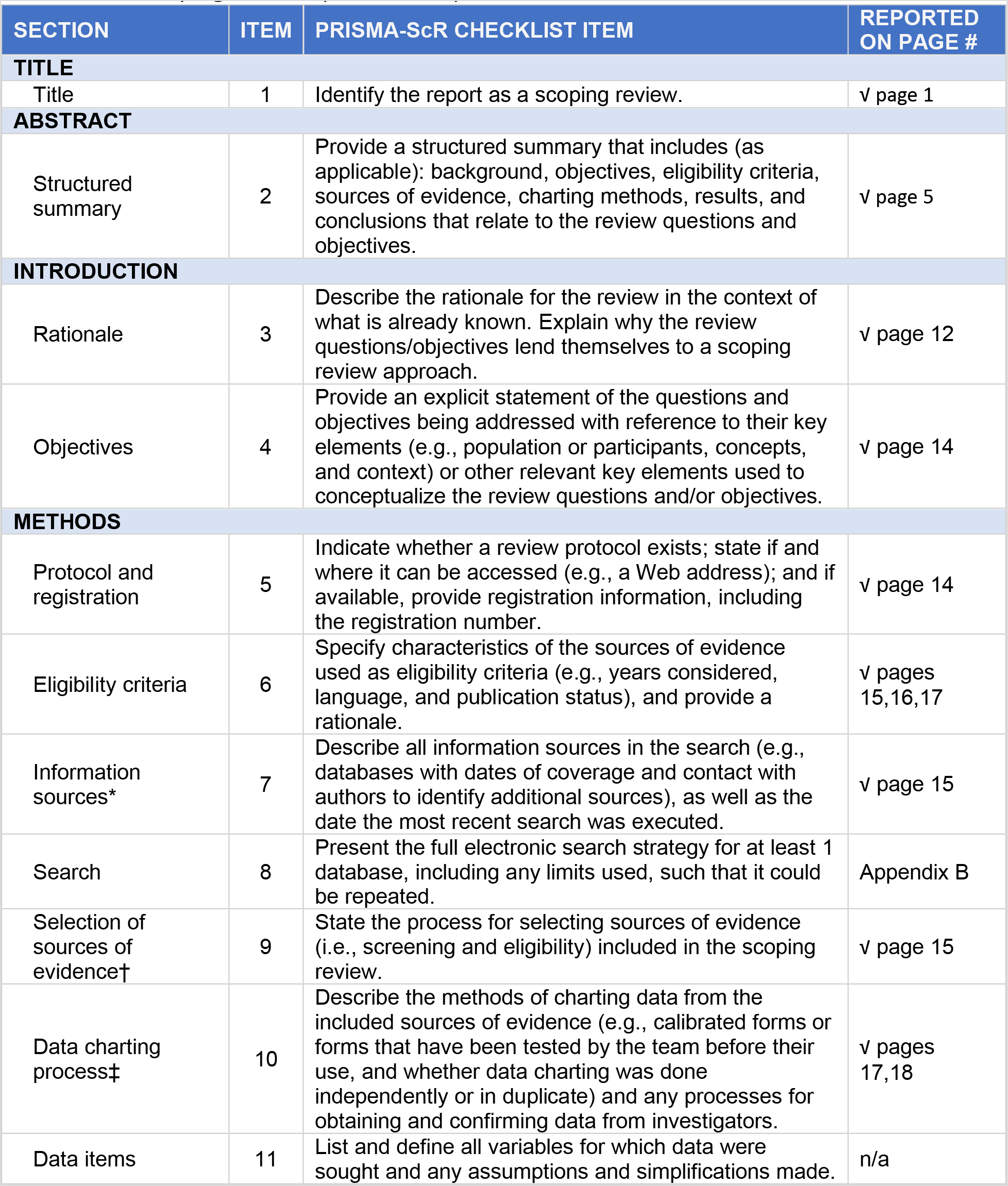
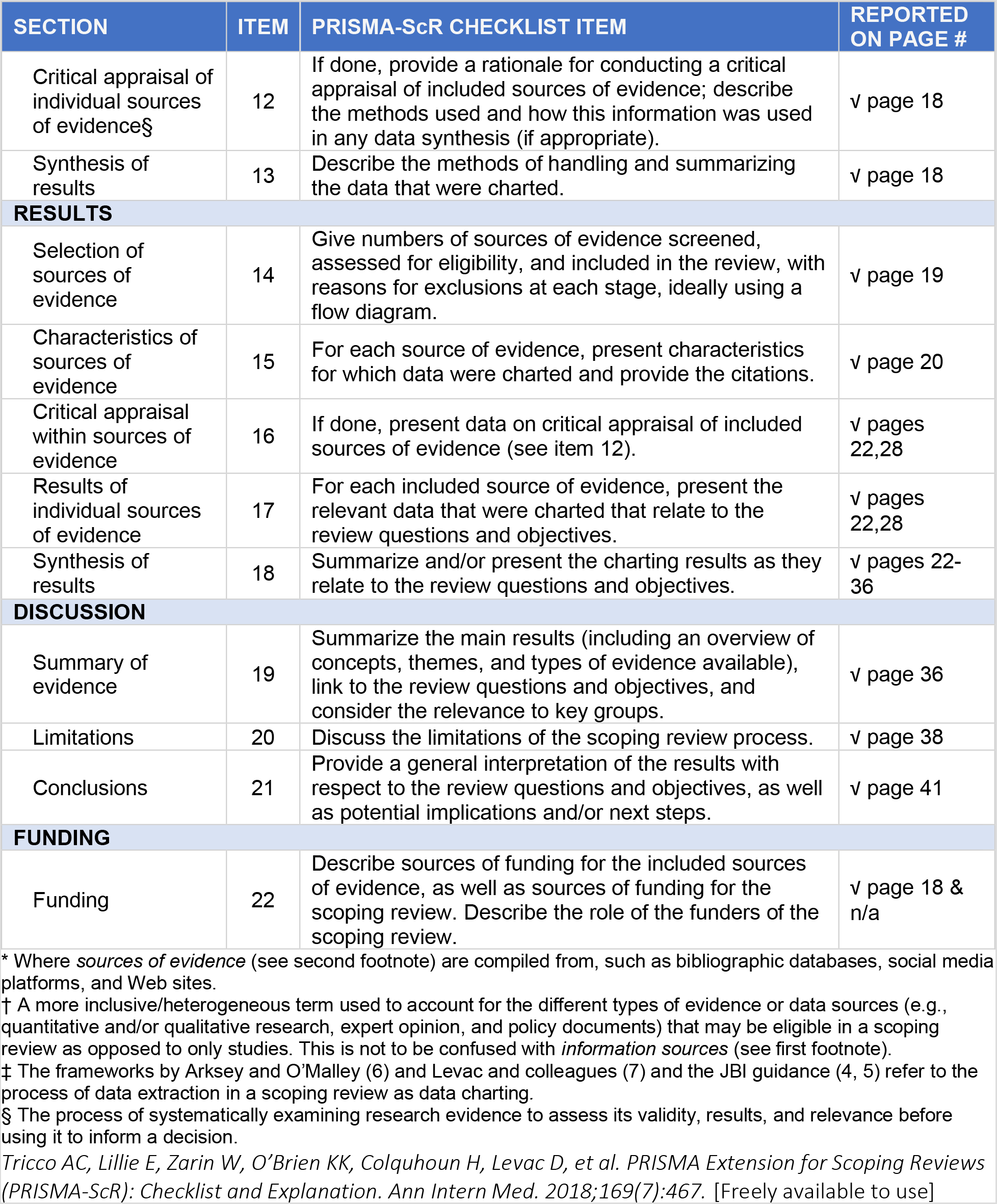

## APPENDIX B: Search strategy for MEDLINE (Ovid)

1. exp Transgender Persons/
2. exp Gender Dysphoria/
3. exp Transsexualism/
4. 1 OR 2 OR 3
5. exp Adolescent/
6. exp Mental Disorders/
7. exp Mental Health/
8. 6 OR 7
9. (Transgender* or Trans or Transsexual* or Travesti* or Gender identity disorder or Gender non-conform* or Gender dysphoria or Gender incongruen*).mp.
10. 4 OR 9
11. (Adolescen* or Teen* or Teenager* or Teenage girl or Teenage boy or Youth).mp.
12. 5 OR 11
13. (Mental health or Mental wellbeing or Mental distress or Mental illness or Psychopathology or Mood disorder or Anxiety* or Depressi* or Substance abuse or Alcohol abuse or Suicid* or Self-harm or Eating disorder or Autism spectrum disorder or Asperger*).mp.
14. 8 OR 13
15. 10 AND 12 AND 14
16. limit 15 to (Afrikaans or Dutch or English or French or Spanish)

## APPENDIX C: Quality appraisal tool used for prevalence studies

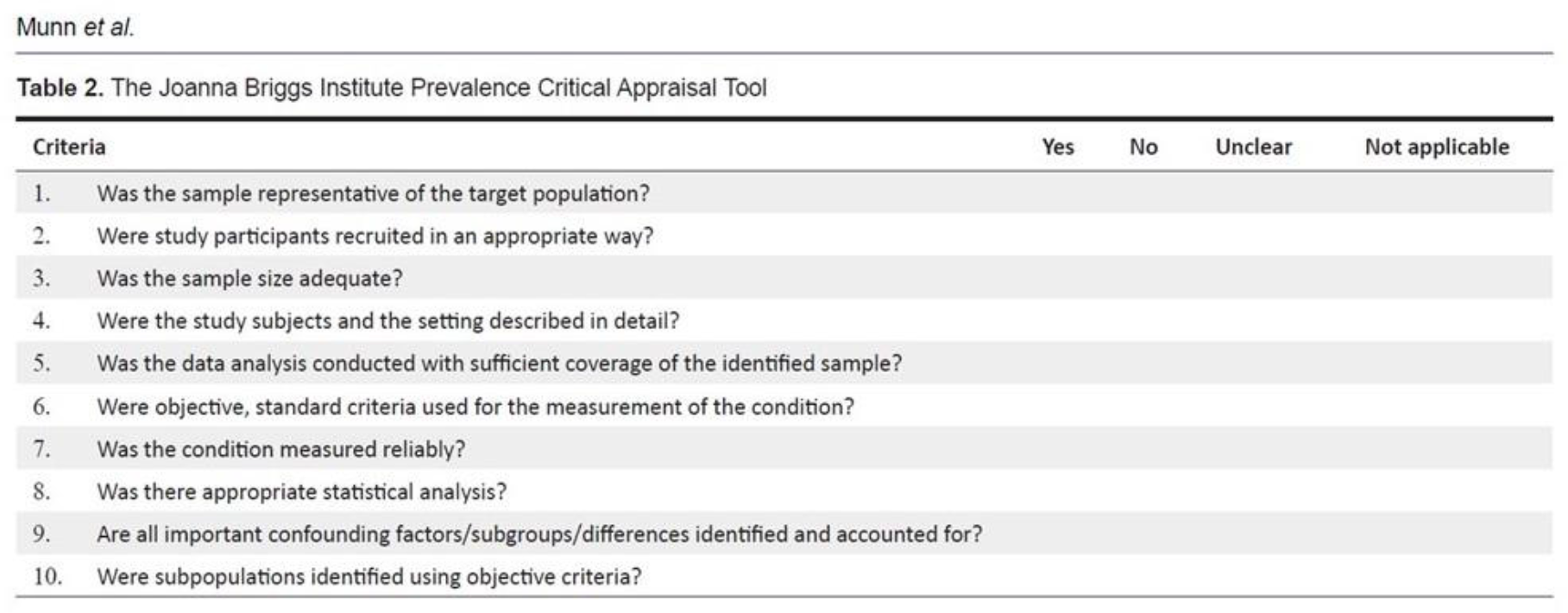

[Table 2 from (reference 50) Munn Z, Moola S, Riitano D, Lisy K. The Development of a Critical Appraisal Tool for Use in Systematic Reviews: Addressing Questions of Prevalence. Int J Health Policy Manag. 2014;3(3):123–8]

